# Correlation of age and the diameter of the cervical nerve roots C5 and C6 during the first two years of life analysed by high-resolution ultrasound imaging

**DOI:** 10.1101/2022.01.26.22269461

**Authors:** Jacoba van der Linde, Carole Jenny, Thomas Hundsberger, Philip J. Broser

## Abstract

**Aim:** To analyse the increase in diameter of the nerve roots C5 and C6 in early childhood.

**Methods:** The nerve roots of 56 children subjects aged 0 days to 10 years (47 younger than 2 years) were examined by high-resolution ultrasound imaging. The correlation of diameter and age was statistically tested and a logarithmic regression analysis was performed to develop a logarithmic growth model.

**Results:** The increase in nerve root diameter is greatest during the first two years of life and then the growth rate decreases steadily. The relationship between age and diameter follows a logarithmic curve (p < 10^−8^).

**Interpretation:** The main increase in the diameter of the nerve roots happens in the first two years of life. Comparing data from a previous study, our data also suggest that the maturation of the proximal part of the median nerve is comparable to the maturation of its distal segments. This suggests a synchronous maturation of the axons and myelin sheath for the whole extent of the nerve, from the radix to its very distal part.

**What this paper adds:** Normative values for the size of the cervical nerve roots C5 and C6, an insight into the maturation of the proximal parts of the peripheral nervous system, and the correlation between age and cervical root diameter.

**Highlights:**

- Maturation of the nerve roots C5 and C6 in children from 0 to two years of age.
- Reference values from the diameter of the C5 and C6 nerve roots of children up to two years.

## 1. INTRODUCTION

An increase in nerve conduction velocity is a crucial hallmark of peripheral nervous system maturation in children during the first years of life [Raimbault, 1998]. Besides a basic understanding of the maturation of Schwann cells and the development of the nerve sheath [Kaplan, 2006], non-invasive observation of in vivo morphological changes was not possible until the introduction of dynamic high-resolution ultrasound imaging of the peripheral nervous system [Walker, 2011]. *In vivo* monitoring of the maturation of the peripheral nervous system allows understanding of the underlying physiology and provides diagnostic and therapeutic knowledge to manage pathophysiological situations like nerve injuries and plexus palsies.

A previous study [Jenny, 2020] demonstrated that the cross-sectional area (CSA) of the median nerve increases in a logarithmic manner from birth up to the age of ten years. Interestingly, the nerve conduction velocity also increases in the same logarithmic manner [García-García, 2004; Parano, 1993]. However, at present it is unclear if the proximal parts of the peripheral nervous system, i.e., nerve roots, develop in a similar fashion as the distal parts of the peripheral nervous system. Wang et al. [Wang, 2020] recently published normative values for the size of nerve roots in children; however, they categorised their patients in age groups of 1-3 years, so a year-wise detailed analysis of the growth rate is not possible. Therefore, we conducted this study to analyse the growth rate of cervical roots during the first two years of life in detail.

## 2. MATERIALS AND METHODS

Children between 0 to 10 years of age admitted to the Children’s Hospital of Eastern Switzerland between February 2021 and September 2021 were recruited for this study. The children were hospitalized due to a mild trauma, such as a radius fracture, or mild non— chronic diseases (e.g., human respiratory syncytial virus infections). Our target sample size was 50 children, with the majority belonging to the 0-2 years age group. All children older than 2 years were assigned to the comparison group to compare our results with the existing values in the literature [Druzhinin, 2019]. Children younger than 2 years were further divided into subgroups based on increments of 100 days of age with at least three children in each group. Criteria for exclusion were a premature birth, family history of neurological diseases, and chronic or severe acute diseases. The study was approved by the Institutional Ethics Committee (EKOS, approval no: EKSG 19/166). Caregivers of all participants signed the informed consent form for including the child into the study.

For the ultrasound examination, a Canon Aplio i800 machine (Canon Medical Systems, Tokyo, Japan) with two ultrasound probes, i18LX5 (max scanning frequency of 18 MHz) for older children and i22LH8 (max scanning frequency of 22 MHz) for younger children, was used. As most of the children examined were below 2 years, the i22LH8 was used more often. The ultrasound device settings were the same as reported by [Jenny, 2020]. The‘general’ setting, with an imaging depth of 30 mm, was used for the i18LX5 probe, and the‘general’ setting, with an imaging depth of 17.5 mm, was used for the i22LH8 probe (footprint of 26 mm). This gave an axial resolution in the range of 100 μm for the i18LX5 probe and 50 μm for the i22LH8 probe [Jenny, 2020].

For the ultrasound examination, the participant’s head was tilted slightly to the opposite side and the ultrasound probe was then placed at the lower part of the neck (see Figure 1). To identify the nerve roots, the authors first looked for the transverse process of the C7 cervical vertebra [Lapegue, 2014; Peer, 2013; Gruber, 2018] which is recognised by its‘bull’s horn’ appearance. Once the‘bulls’ horn’ was identified, the ultrasound probe was moved one vertebra up cranially to scan for the nerve root C6. After examining the C6, the probe was moved one vertebra further cranially to scan for the nerve root C5. If possible, both the left and right sides of the neck were examined.

**Figure 1.**
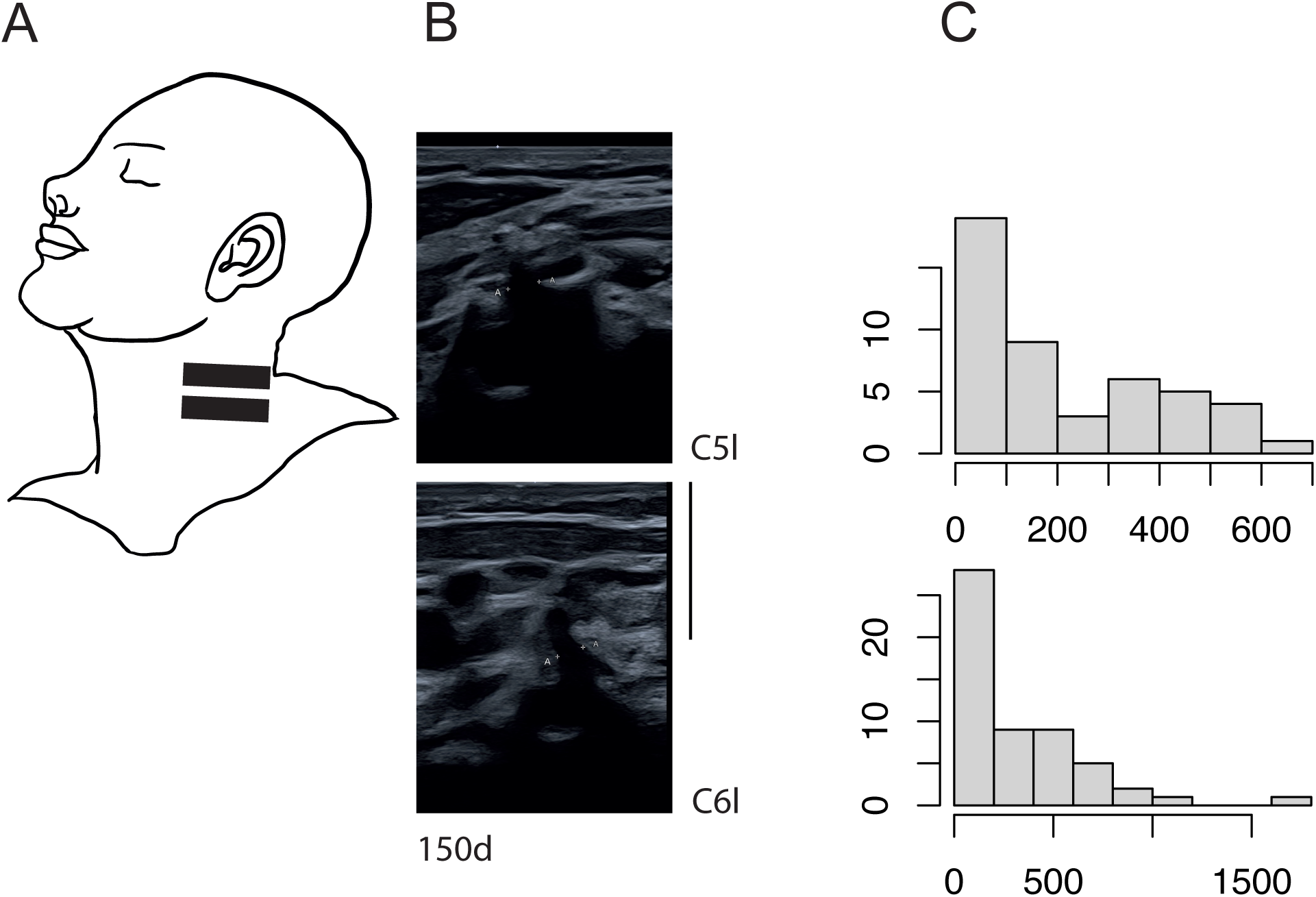
(A) Placement of the ultrasound probe on the neck to scan for the nerve roots C6 and C5. (B) Example images of the two nerve roots C5-left and C6-left at the age of 150 days. Two points «A» limit the diameter. (Diameter C5: 1.71mm, C6: 1.74mm, scale bar applies to both images and is 10mm) (C) Age distribution of all subjects included in the study. Top image: Age distribution of the subjects in the main study group (aged < 2 years, precise age < 700 days). Each age segment represents 100 days.

During the examination, videos of the examined nerve roots were taken to compensate for the incompliance in the study population. After the examination from the video a still image of the nerve root was selected and the diameter of the nerve root was measured just distal from the foramen intervertebrale before the root bifurcates into different branches. The data was analysed using the R software [R core team, 2021].

A Mann-Whitney U test was used to compare the cross-sectional diameter on the left and right side. To put the age and diameter in correlation and test for statistical significance, Kendall’s correlation test was used.

To avoid interobserver reliability, a single examiner, the author JVDL, educated all participants, prepared, and examined them to take the measurements. Intraobserver reliability was also tested by repeating the same measurements on the ultrasound images of three subjects on three different days.

## 3. RESULTS

A total of 61 patients were screened to obtain the target sample size. Five children had to be excluded from the study – two had injuries to the median nerve, one to the ulnar nerve, one showed signs of an inflammatory neuropathy, and one due to caregivers withdrawal from the study. Thus, 56 children (25 females) aged between 9 days and 10 years of age were included in the study, of which, 47 belonged to < 2 years age group and were included in the main study group. The age distribution of the whole study group is shown in Figure 1 - Panel C1, and that of the children aged below 2 years is given in Figure 1 - Panel C2.

### Imaging of the roots C5 and C6

Figure 2 shows ultrasound images of the cervical roots C5 and C6 for children aged 20, 50, and 500 days (16.4 months) in age. The figure qualitatively shows the increase in diameter of the root from birth to the age of one and a half years.

**Figure 2.**
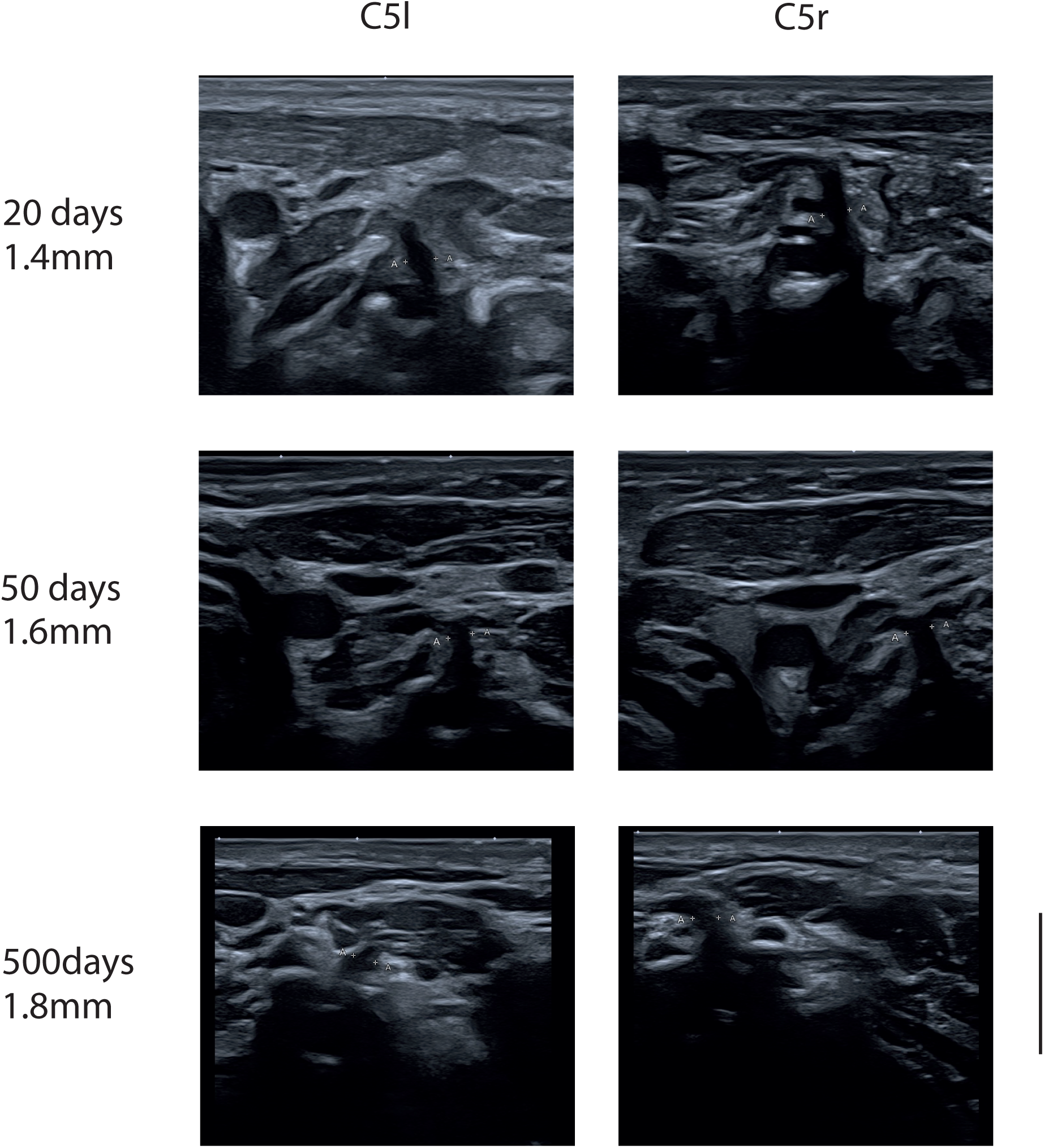
Typical ultrasound images of the nerve roots C5-left and C5-right for all three locations from three participants aged 26, 50, and 500 days. The scale bar on the lower right corner is 10mm and applies to all images.

### Cross-sectional diameter

The obtained measurements of the diameter of the nerve roots are listed in Table 1. The Mann-Whitney U test was used to compare the left and right sides; no statistically significant difference was found between the two sides (median difference for root C5: 0.04 mm; median difference for root C6: 0.03 mm). Visual inspection of the ultrasound still images revealed an increase in the size of the nerve roots with age (see Figure 2). To visualize this qualitative finding, scatter plots depicting the nerve root diameters at different ages are shown in Figure 3 for the complete study and in Figure 4 for the main study group (children < 2 years). The Kendall correlation test was applied for all four roots (C6-r, C6-l, C5-r, C5-l) to test the correlation between age and diameter [Hollander, 1973]. The test showed highly significant correlations for all four locations (C6-r: tau=0.52, p < 10^−7^; C6-l: tau=0.52, p < 10^−7^; C5-r: tau=0.53, p < 10^−7^; C5-l: tau=0.50, p < 10^−6^), confirming a statistically significant increase in the diameter with age for all four locations.

**Table 1.** Demographics and cervical root measurements

| Demographics |  |  |  | Diameters in [mm] |  |  |  |
| --- | --- | --- | --- | --- | --- | --- | --- |
| age[d] | weight[kg] | length[cm] | sex | C6r | C6l | C5r | C5l |
| 9 | 3.3 | 49 | m | 1.53 | 1.55 | 1.40 | 1.54 |
| 14 | 3.7 | 51 | f | 1.42 | 1.34 | 1.40 | 1.37 |
| 16 | 4.7 | 55 | f | 1.60 | 1.37 | 1.55 | 1.51 |
| 25 | 4.4 | 53 | m | 1.44 | 1.53 | 1.44 | 1.48 |
| 27 | 3.8 | 53 | m | 1.42 | 1.42 | 1.19 | 1.19 |
| 38 | 3.7 | 51 | f | 1.54 | 1.54 | 1.52 | 1.57 |
| 39 | 5.5 | 57 | m | 1.55 | 1.52 | 1.57 | — |
| 40 | 3.8 | 56 | f | 1.52 | 1.52 | 1.21 | 1.47 |
| 41 | 4.2 | 53 | f | 1.63 | 1.63 | 1.53 | 1.53 |
| 47 | 4.8 | 52 | m | 1.24 | 1.31 | 1.37 | 1.30 |
| 53 | 5.2 | 54 | f | 1.50 | 1.50 | 1.50 | 1.50 |
| 55 | 3.8 | 56 | m | 1.53 | 1.61 | — | 1.44 |
| 60 | 5.4 | 60 | m | 1.50 | — | 1.70 | — |
| 63 | 5.1 | 60 | f | 1.53 | 1.55 | 1.45 | 1.46 |
| 68 | 4.8 | 57 | — | 1.55 | 1.59 | 1.60 | 1.46 |
| 70 | 5.6 | 59 | f | 1.55 | 1.56 | 1.42 | 1.46 |
| 78 | 4.5 | 58 | f | 1.47 | 1.50 | 1.50 | 1.60 |
| 82 | 5.6 | 64 | m | 1.56 | 1.58 | 1.54 | 1.56 |
| 93 | — | — | f | 1.60 | 1.60 | 1.50 | 1.70 |
| 107 | 6.5 | 61 | m | 1.89 | 1.79 | 1.73 | 1.89 |
| 110 | 6.1 | 63 | m | 1.53 | 1.43 | 1.52 | 1.45 |
| 123 | 3.8 | 49 | f | 1.18 | 1.26 | 1.08 | 1.26 |
| 126 | 6.5 | 64 | m | 1.44 | 1.40 | 1.40 | 1.43 |
| 133 | 7.7 | 65 | m | 1.63 | 1.74 | 1.70 | 1.63 |
| 135 | 6.2 | 62 | f | 1.55 | 1.54 | 1.53 | 1.50 |
| 141 | 8.1 | 68 | f | 1.70 | — | 1.70 | — |
| 146 | 8.4 | 64 | m | 1.70 | 1.70 | 1.60 | 1.60 |
| 153 | 7.4 | 67 | m | 1.74 | 1.76 | 1.86 | 1.71 |
| 227 | 9.8 | 72 | m | 1.76 | 1.57 | 1.79 | 1.83 |
| 251 | 6.1 | 64 | f | 1.68 | 1.71 | 1.66 | 1.70 |
| 257 | 8.6 | 75 | m | 1.71 | 1.70 | 1.60 | 1.87 |
| 306 | 7.4 | 70 | f | 1.80 | 1.90 | 1.70 | 1.70 |
| 320 | 9.1 | — | m | 1.60 | 1.60 | 1.65 | 1.60 |
| 322 | 7.1 | 69 | f | 1.59 | — | 1.60 | — |

Table 1: Cervival Root Data
| Demographics |  |  |  | Diameters in [mm] |  |  |  |
| --- | --- | --- | --- | --- | --- | --- | --- |
| age[d] | weight[kg] | length[cm] | sex | C6r | C6l | C5r | C5l |
| 325 | 8.8 | 79 | m | 1.83 | 1.78 | 1.74 | 1.78 |
| 335 | 8.2 | 70 | m | 1.73 | 1.66 | 1.72 | 1.70 |
| 336 | 10.5 | 74 | m | 1.80 | 1.80 | 1.80 | 1.70 |
| 401 | 11.0 | 77 | m | 1.71 | 1.73 | 1.70 | 1.67 |
| 417 | 11.6 | 79 | m | 1.70 | 1.80 | 1.80 | 1.70 |
| 420 | 10.7 | 76 | f | 1.77 | 1.78 | 1.75 | 1.76 |
| 473 | 19.0 | — | m | — | 1.78 | — | 1.75 |
| 488 | 12.3 | 80 | m | 1.64 | 1.60 | 1.63 | — |
| 509 | 10.7 | 79 | f | — | 1.80 | — | 1.70 |
| 584 | — | — | m | 1.65 | 1.64 | 1.56 | 1.54 |
| 590 | 9.9 | — | m | 1.82 | 1.83 | 1.72 | 1.78 |
| 598 | 12.7 | 85 | f | 1.71 | 1.83 | 1.92 | 1.87 |
| 670 | 12.0 | 86 | m | 1.72 | 1.70 | 1.63 | 1.68 |
| 736 | 11.3 | 85 | f | 1.76 | 1.79 | 1.72 | 1.75 |
| 750 | 12.2 | 85 | f | — | 1.74 | — | 1.77 |
| 772 | 11.8 | 86 | m | 2.00 | 2.10 | 2.00 | 2.00 |
| 781 | 13.0 | 88 | f | 2.10 | 2.00 | 2.10 | — |
| 829 | 10.5 | 86 | f | 1.80 | 1.80 | 1.73 | 1.80 |
| 960 | 14.5 | — | m | 1.87 | 1.75 | 1.71 | 1.76 |
| 1,039 | 12.8 | — | m | 1.60 | 1.60 | 1.60 | 1.60 |
| 1,623 | 17.0 | — | f | 2.00 | 2.00 | 1.70 | 1.90 |
| 3,753 | 32.9 | 144 | m | 2.30 | 2.50 | 2.90 | — |

**Table 2.** Median values of the nerve roots. The median, 25^th^, and 75^th^ quartiles are listed for all nerve roots - C6-right, C6-left, C5-right, and C5-left. Age segments are of 120 days

| Age |  | C6r |  |  | C6l |  |  | C5r |  |  | C5l |  |  |
| --- | --- | --- | --- | --- | --- | --- | --- | --- | --- | --- | --- | --- | --- |
| age from [d] | age to [d] | Q25 | Median | Q75 | Q25 | Median | Q75 | Q25 | Median | Q75 | Q25 | Median | Q75 |
| 0 | 120 | 1.48 | **1.53** | 1.55 | 1.50 | **1.53** | 1.56 | 1.40 | **1.50** | 1.54 | 1.46 | **1.48** | 1.53 |
| 120 | 240 | 1.53 | **1.61** | 1.70 | 1.43 | **1.60** | 1.74 | 1.50 | **1.56** | 1.70 | 1.45 | **1.60** | 1.70 |
| 240 | 360 | 1.69 | **1.71** | 1.73 | 1.64 | **1.70** | 1.71 | 1.63 | **1.66** | 1.73 | 1.77 | **1.83** | 1.85 |
| 360 | 480 | 1.63 | **1.77** | 1.80 | 1.66 | **1.78** | 1.80 | 1.66 | **1.71** | 1.73 | 1.70 | **1.70** | 1.70 |
| 480 | 600 | 1.71 | **1.71** | 1.74 | 1.75 | **1.78** | 1.79 | 1.73 | **1.75** | 1.77 | 1.69 | **1.70** | 1.73 |
| 600 | 720 | 1.64 | **1.64** | 1.64 | 1.69 | **1.78** | 1.79 | 1.63 | **1.63** | 1.63 | 1.71 | **1.73** | 1.74 |
| 720 | 840 | 1.68 | **1.71** | 1.77 | 1.73 | **1.83** | 1.83 | 1.64 | **1.72** | 1.82 | 1.66 | **1.78** | 1.83 |
| 840 | 960 | 1.72 | **1.72** | 1.72 | 1.70 | **1.70** | 1.70 | 1.63 | **1.63** | 1.63 | 1.68 | **1.68** | 1.68 |
| 960 | 1080 | 1.88 | **2.00** | 2.05 | 1.78 | **1.90** | 2.02 | 1.86 | **2.00** | 2.05 | 1.76 | **1.77** | 1.89 |

**Figure 3.**
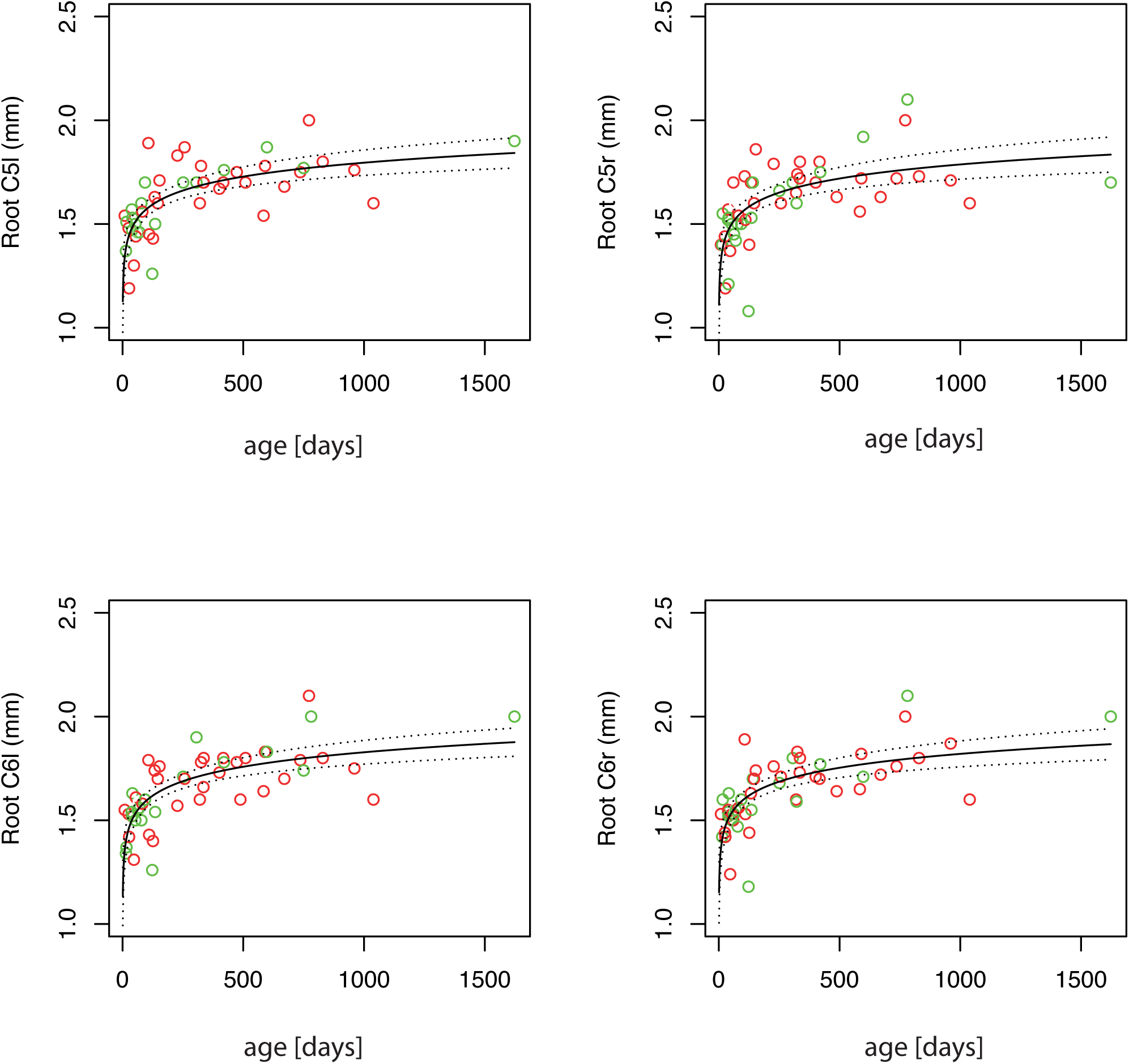
Top left: Root C5-left; Top right: Root C5-right; Bottom left: Root C6-left; Bottom right: Root C6-right. The panels cover the whole age range of the study (main study group and comparison group). The measured diameter is plotted against age (green dots: right arm; red dots: left arm). The logarithmic regression curve is plotted in each panel as a black line, with the upper and lower limits of the 95% confidence intervals plotted as dotted lines above and below.

**Figure 4.**
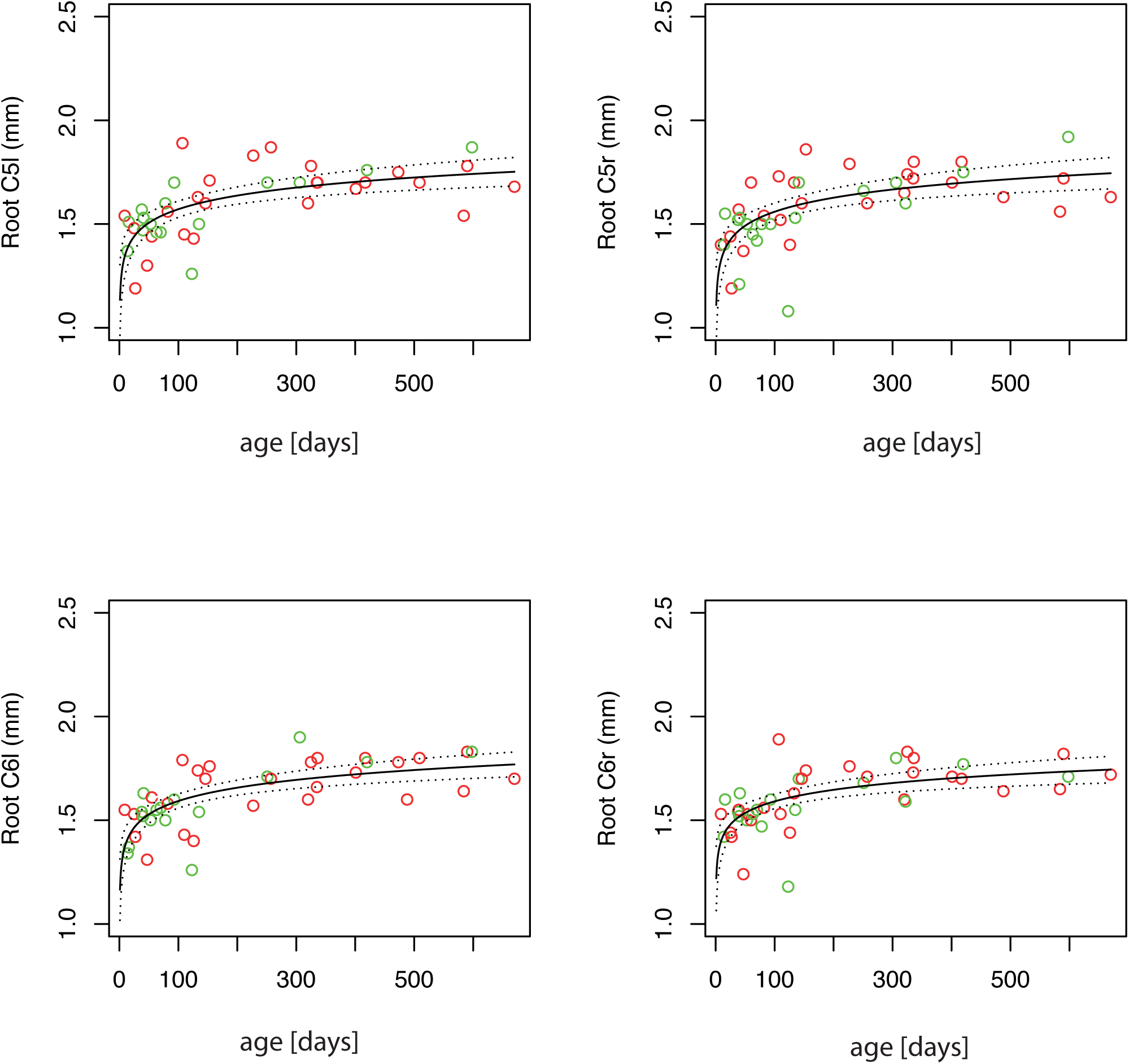
Top left: Root C5-left; Top right: Root C5-right; Bottom left: Root C6-left; Bottom right: Root C6-right. The panels focus on the main study group. The measured diameter is plotted against age (green dots: right arm; red dots: left arm). The logarithmic regression curve is plotted in each panel as a black line, with the upper and lower limits of the 95% confidence intervals plotted as dotted lines above and below.

### Logarithmic Regression Curve

The increase in diameter was most significant during the first two years of age, and the slope of increase flattened with increasing age. This is typical of processes that are governed by a logarithmic model. Therefore—as previously reported [Jenny, 2020]—a logarithmic model:

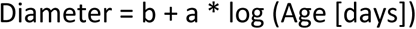

was chosen, where‘a’ is the slope and‘b’ is the intersection, and a regression analysis, as described by Natale and Rajagopalan [Natale, 2014], was performed. The model was tested for each location and for the whole study group and the main study group, and‘a’ and‘b’ were numerically determined by regression analysis [Wilkinson, 1973]. The parameters obtained for the whole study group were as follows: (main study group and control group together), C6-r: a=0.10, b=1.15, Standard error of the regression=0.128, p < 10^−8^; C6-l: a=0.10, b=1.13, Standard error of the regression=0.123, p < 10^−9^; C5-l: a=0.10, b=1.11, Standard error of the regression=0.146, p < 10^−7^; C5-r: a=0.10, b=1.13, Standard error of the regression=0.126, p < 10^−8^.

The values obtained focusing on the main study group were as follows: C6-r: a=0.08, b=1.22, Standard error of the regression=0.118, p < 10^−6^; C6-l: a=0.09, b=1.17, Standard error of the regression=0.114, p < 10^−7^; C5-l: a=0.10, b=1.11, Standard error of the regression=0.139, p < 10^−6^; C5-r: a=0.10, b=1.34, Standard error of the regression=0.127, p < 10^−6^.

The results of the logarithmic model are plotted in Figures 3 and 4. In summary, the regression analysis showed a highly significant result with a low standard error of the regression, suggesting that the increase in diameter is well described by the logarithmic model.

## 4. Discussion

This study aimed to evaluate the relationship between the diameter of the cervical roots C5 and C6 and age in children ranging from neonates to 10 years of age. No differences were observed in the nerve root diameter between the left and right sides or with gender differences. However, we observed a statistically highly significant correlation between age and nerve-root diameter. Similar to the previously reported results for the median nerve [Jenny, 2020], we obtained a logarithmic relation between age and cross-sectional diameter of the nerve roots, i.e., the increase in the diameter of the C5 and C6 nerve roots follows a logarithmic growth curve. Notably, the most significant increase in the diameter happened during the first 24 months of life, and from then on, the increment slows down significantly. Jenny described that the cross-sectional area of the median nerve follows a logarithmic growth curve during the first two years of life, and further proposed a close correlation between nerve conduction velocity and nerve size. After the evidence that the increase in nerve conduction velocity follows the same logarithmic curve as the increase in area, the authors further questioned how and when does the proximal part of the nerve mature.This study now shows that the maturation of the nerve roots follows the same temporal pattern found for the median nerve [Jenny, 2020]. Both studies together suggests a synchronous maturation of the axons and myelin sheath throughout the whole nerve, from the radix to the very distal part of a nerve.

However, despite the substantial findings of our research, this study raises further questions. First, is the functional maturation (increase in nerve conduction velocity) of the proximal parts of the nerve like the one that is occurring at the distal parts? Second, is the maturation of the lower body nerves like the maturation of the median nerve and the other nerves of the upper extremities? These questions will be addressed in future studies.

A limitation of the study is the small number of subjects analysed, especially those belonging to the two to ten years age group (n=10). Also, this study was designed as a cross-sectional study; a longitudinal approach would be more appropriate to address the question of increase in the nerve diameter in different individuals. Further studies should aim to involving a larger sample size studies that is more representative of the study population and study them over longer periods.

## Data Availability

All data produced in the present work are contained in the manuscript.

## Abbreviations

C5-l: cervical nerve root 5 on the left side
C5-r: cervical nerve root 5 on the right side
C6-l: cervical nerve root 6 on the left side
C6-r: cervical nerve root 6 on the right side

## Acknowledgments

We thank the children and parents who participated in the study. This study was supported by a grant from the Science Commission of the Cantonal Hospital, St. Gallen (20/31)

## Conflicts of Interest

The authors declare no conflicts of interest.

## Figure legends

**CONSORT diagram:** CONSORT (2010) flowchart of study selection.

## Notes

### Competing Interest Statement

The authors have declared no competing interest.

### Funding Statement

This study was funded by Kantonsspital St Gallen.

### Author Declarations

Ethikkommission Ostschweiz EKOS Web: https://www.sg.ch/gesundheit-soziales/gesundheit/gremien.html Project approval no: EKSG 19/166

